# Modelling the Anatomical Distribution of Neurological Events in COVID-19 Patients: A Systematic Review

**DOI:** 10.1101/2020.10.21.20215640

**Authors:** Nicholas Parsons, Athanasia Outsikas, Annie Parish, Rebecca Clohesy, Nilam Thakkar, Fiore D’Aprano, Fidel Toomey, Shailesh Advani, Govinda Poudel

## Abstract

**Background:** Neuropathology caused by the coronavirus disease 2019 (COVID-19) has been reported across several studies. The characterisation of the spatial distribution of these pathology remains critical to assess long and short-term neurological *sequelae* of COVID-19. To this end, Mathematical models can be used to characterise the location and aetiologies underlying COVID-19-related neuropathology.

**Method:** We performed a systematic review of the literature to quantify the locations of small neurological events identified with magnetic resonance imaging (MRI) among COVID-19 patients. Neurological events were localised into the Desikan-Killiany grey and white matter atlases. A mathematical network diffusion model was then used to test whether the spatial distribution of neurological events could be explained via a linear spread through the structural connectome of the brain.

**Findings:** We identified 35 articles consisting of 123 patients that assessed the spatial distribution of small neurological events among COVID-19 patients. Of these, 91 patients had grey matter changes, 95 patients had white matter changes and 72 patients had confirmed cerebral microbleeds. White matter events were observed within 14 of 42 white matter bundles from the IIT atlas. The highest proportions (26%) of events were observed within the bilateral corticospinal tracts. The splenium and middle of the corpus callosum were affected in 14% and 9% of the cases respectively. Grey matter events were spatially distributed in the 41 brain regions within the Desikan-Killiany atlas. The highest proportions (∼10%) of the events were observed in areas including the bilateral superior temporal, precentral, and lateral occipital cortices. Sub-cortical events were most frequently identified in the Pallidum. The application of a mathematical network diffusion model suggested that the spatial pattern of the small neurological events in COVID-19 can be modelled with a linear diffusion of spread from epicentres in the bilateral cerebellum and basal ganglia (Pearson’s *r*=0.41, *p*<0.001, corrected).

**Interpretation:** To our knowledge, this is the first study to systematically characterise the spatial distribution of small neurological events in COVID-19 patients and test whether the spatial distribution of these events can be explained by a linear diffusion spread model. The location of neurological events is consistent with commonly identified neurological symptoms including alterations in conscious state among COVID-19 patients that require brain imaging. Given the prevalence and severity of these manifestations, clinicians should carefully monitor neurological symptoms within COVID-19 patients and their potential long-term *sequelae*.

---

The coronavirus disease 2019 (COVID-19) is caused by severe acute respiratory syndrome coronavirus 2 (SARS-CoV-2).^1^ Typically, COVID-19 patients present with fever, cough, fatigue, and dyspnoea, with approximately 20% of cases developing severe life-threatening disease.^1,2^ Extrapulmonary symptoms are also being reported including altered conscious state, seizures and focal neurological injuries, raising concerns over the long-term neurological *sequelae* of COVID-19.^3-8^ As of October 15^th^ 2020, more than 38.4 M cases and 1.1 M deaths have been reported globally, with cases rising rapidly in the U.S., India and Brazil.^9^

Several neuroimaging studies have suggested that neurological symptoms in COVID-19 patients are linked to a broad range of acute neurological events, particularly those of cerebrovascular nature. A recent systematic review has shown that these events range from large ischemic strokes to small and localised haemorrhages, vascular thrombosis and microbleeds^10^. The presence of cerebral microbleeds (small 2–5 mm perivascular hemosiderin deposits) are also being reported and are presumed to be features of small vessel disease. These smaller neurological events can manifest as fluid-attenuated inversion recovery (FLAIR) signal abnormalities in either grey or white matter,^10^ or localised signal changes as measured with T1-weighted, susceptibility-weighted, and diffusion-weighted MRI.^11-13^ These events are being associated with increased risk of stroke and poorer functional recovery, which may compromise long-term cognitive outcomes,^11,13-15^ and may therefore reflect a selective vulnerability of brain regions to COVID-19.

Emerging evidence suggests COVID-19 patients may display an atypical distribution of neurological events occurring primarily within the corpus callosum and deep subcortical structures.^7,16-18^ However, these distributions are not well understood and may benefit from mathematical modelling in order to characterise the pattern of distribution and potential epicentres of spread. For instance, network diffusion models (NDM) can emulate the pattern of pathological spread in the brain via white matter pathways, and have been useful in modelling the spread of disease in other neurological conditions such as Huntington’s and Alzheimer’s disease.^19-21^ However, to date there has been no application of any mathematical model to assess and map the distribution of neurological events associated with COVID-19.

This systematic review aims to (1) critically review current literature investigating the location of neurological events in patients with COVID-19 and (2) shed light on the distribution of COVID-19 related neurological events within grey and white matter. Subsequently, we: (1) summarized recent literature on neurological events (2) mapped the spatial distribution of neurological events and (3) utilised NDM to model the spread of neurological events in COVID-19 patients.

## Method

### Protocol registration

This systematic review was registered with the International Prospective Register of Systematic Reviews (PROSPERO: registration number CRD42020201161) and conducted according to PRISMA guidelines.^22^

### Search strategy

We searched Medline, Embase, Scopus and LitCovid databases from 1^st^ January 2020 – 19^th^ July 2020 using the MeSH terms “coronavirus” OR “COVID-19” AND “neurolog*” OR “brain” OR “central nervous system” OR “CNS” AND “MRI” OR “magnetic resonance imaging” OR “hypointensities” OR “microbleeds” OR “cerebral microbleeds” OR “microhemorrhages”. Additional studies were identified by manually searching the reference lists of relevant articles. The search strategy is described in supplementary Fig. 1 in a PRISMA flow-chart. Search was conducted with help of a health science librarian.

**Fig 1.**
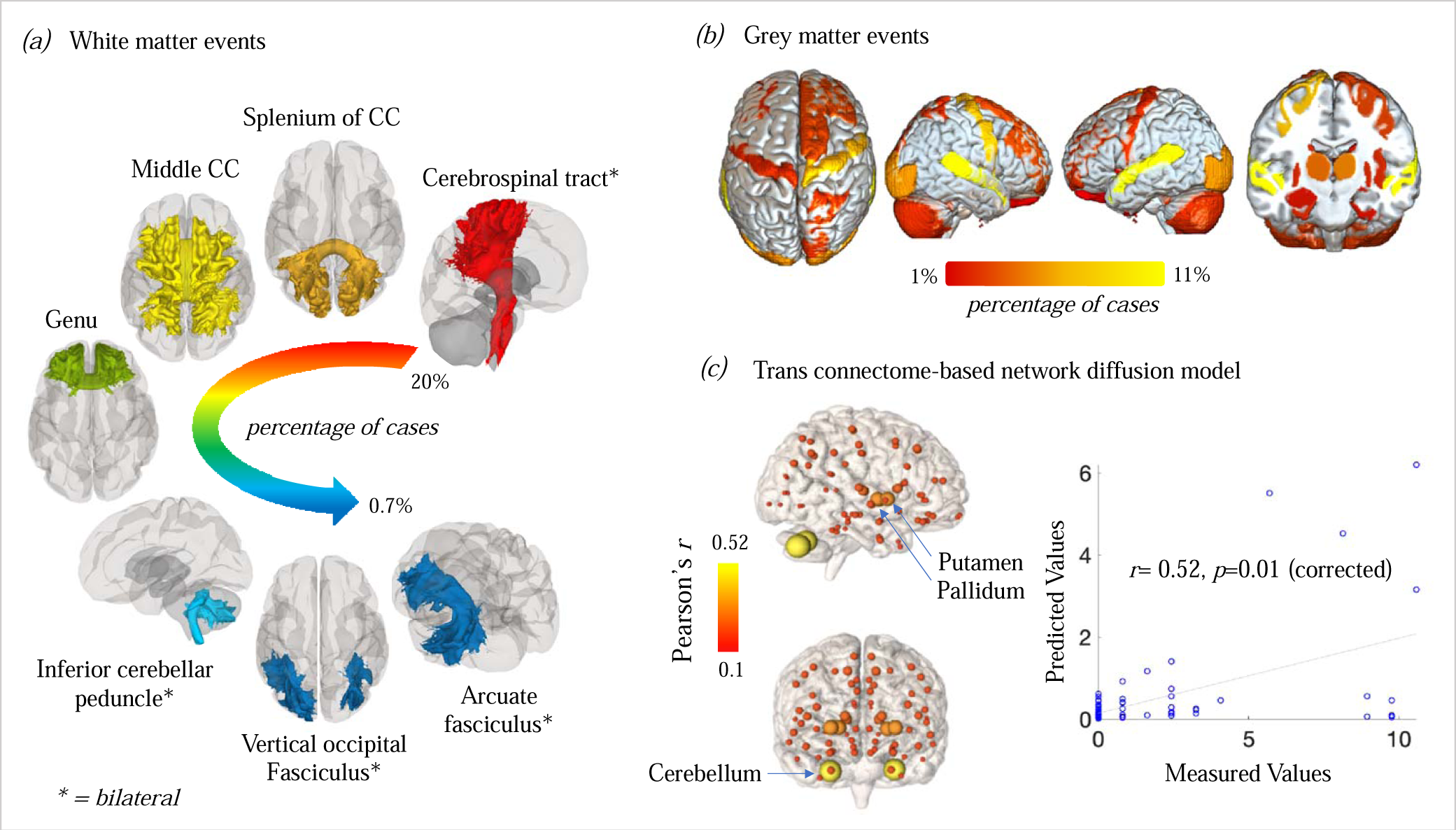
Distribution and modelling of neurological events in the COVID-19 brain.

### Selection criteria

We included case reports, case series and observational studies published in peer-reviewed journals and preprints available in English that identified small neurological events in patients with COVID-19 using MRI. Articles without full texts and studies without laboratory-confirmed COVID-19 patient diagnosis were excluded. Any studies only reporting large cerebrovascular events (such as strokes, infarcts) and diffuse pathology (non-specific) were also excluded.

### Data extraction

Two independent reviewers screened articles by title and abstract for relevance. These studies were then screened for eligibility for inclusion by full text evaluation. For each included article, two independent reviewers extracted data (AP, RC). Disagreements were collaboratively resolved within the team. Instructions detailing the type of information to be extracted and how to record, categorise or code this information was also discussed amongst team members. The following information was extracted from each manuscript: (*a*) country, first author and year of publication; (*b*) sample characteristics (sample size, age group and sex distribution); (*c*) study design; (*d*) clinical symptoms; (*e*) reason for brain imaging; (*f*) type of MRI performed; (*g*) imaging findings; and (*h*) relevant conclusions to assist article interpretation. Two additional reviewers (FD, FT) then validated all the extracted data and the eligibility of each included article.

### Neuroimaging data synthesis and coding

Two expert reviewers (NP, GP) screened each included article to identify the location, distribution, and number of neurological events. These events ranged from microbleeds (observed in SWI or T2* GRE images), white matter hyperintensities (FLAIR images), small lesions, or signal changes in diffusion-weighted MRI within grey or white matter. For each article, events were manually localised to grey or white matter regions based on available MRI images and/or radiological descriptions. The Illinios Institute of Technology (IIT) Desikan-Killiany grey matter atlas incorporating 84 brain regions, was used to label any events located within grey matter.^23^ IIT white matter bundles were used to label any events located within white matter.^23^ FSLeyes neuroimaging software from the FMRIB library was used to visualise white matter tracts and grey matter areas from the IIT atlas.^24^ Any COVID-19 patient with non-specific neuropathological findings (e.g. “juxtacortical white matter”) or without an MRI image or description were excluded from further analyses. This data encoding process generated two tables for grey and white matter regions with columns corresponding to each article and rows corresponding to the name of the region/bundle. Each cell in the table provided information on the number of cases corresponding to the neurological event. A third independent reviewer (FT) validated the encoded data and any discrepancies were discussed and addressed.

### Neuroimaging Data Visualisation

Proportions of events 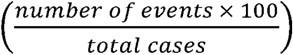 belonging to each encoded region within the IIT Desikan-Killiany grey matter atlas and IIT white matter bundles were used for visualisation. Grey matter events were visualised using MRICroGL software.^25^ White matter events were visualised using MATLAB 2018a and CONN toolbox version 19b. ^26,27^

### Network diffusion model of spread

A graph theoretical meta-analytic model was used to test whether the spatial distribution of small neurological events in the brain can be explained by a spread via the brain’s structural connectome (source code available at: https://github.com/govin2000/covidspread). NDM was used per previous protocols that identified a spatial pattern of pathology in the brain.^19,20^ NDM models the hypothetical distribution of pathology in a brain network (given by a connectome C) over time by linear diffusion, given by: *x*(*t*) = *e*^−*βHt*^ *x*_0_ where *x*_0_ is the initial pattern of the neurological events at *t*= *0*, H is the degree normalised graph Laplacian, and *β* is a diffusivity constant. The unit of the model’s diffusion time *t* is assumed to be days (given the likely progression of 5-14 days^4,8,14^) for the diffusivity constant *β* of 1 per day. *x*(*t*) is a vector of distribution of pathology in the brain when diffusion is seeded from a given region provided by an initial condition *x*_0_. We used a repeated seeding approach, which has previously been used to identify potential epicentres of the spread of neuropathology.^28^ We also used the IIT brain connectome, which is available openly.^23^

NDM generates a vector of distribution of pathology *x*(*t*) over time. We expect that *x*(*t*) should correlate with distribution of neurological events. Thus, Pearson’s correlation coefficient strength and *p*-values were calculated between the empirical proportions of events measured using the systematic review (described above) and *x*(*t*) at all model timepoints (*t*). This process was repeated for all bilateral regions (42 bilateral regions) within the IIT Desikan-Killiany grey matter atlas. The region that showed the largest significant (*p*<0.05, Family-wise error corrected for 84 regions) association with measured neurological events was defined as the seed region.

## Results

The systematic search yielded 461 articles, of which 62 were eligible for full-text assessment (see Supplementary Fig. 1 for PRISMA flow-chart). Of these, 28 were excluded; these were commentaries, response letters and review articles proposing SARS-CoV-2 nervous system invasion but lacking clinical findings. A total of 35 publications reporting small neurological events in patients with COVID-19 were evaluated. Of these, 35 provided specific anatomical detail required for meta-analysis and modelling. These articles contributed 123 unique patients, with a total of 317 neurological events. Of these, 91 patients had grey matter changes, 95 patients had white matter changes and 72 patients had confirmed cerebral microbleeds. Further details on the presentation of grey and white matter events can be found in table 1 below.

**Table 1.**
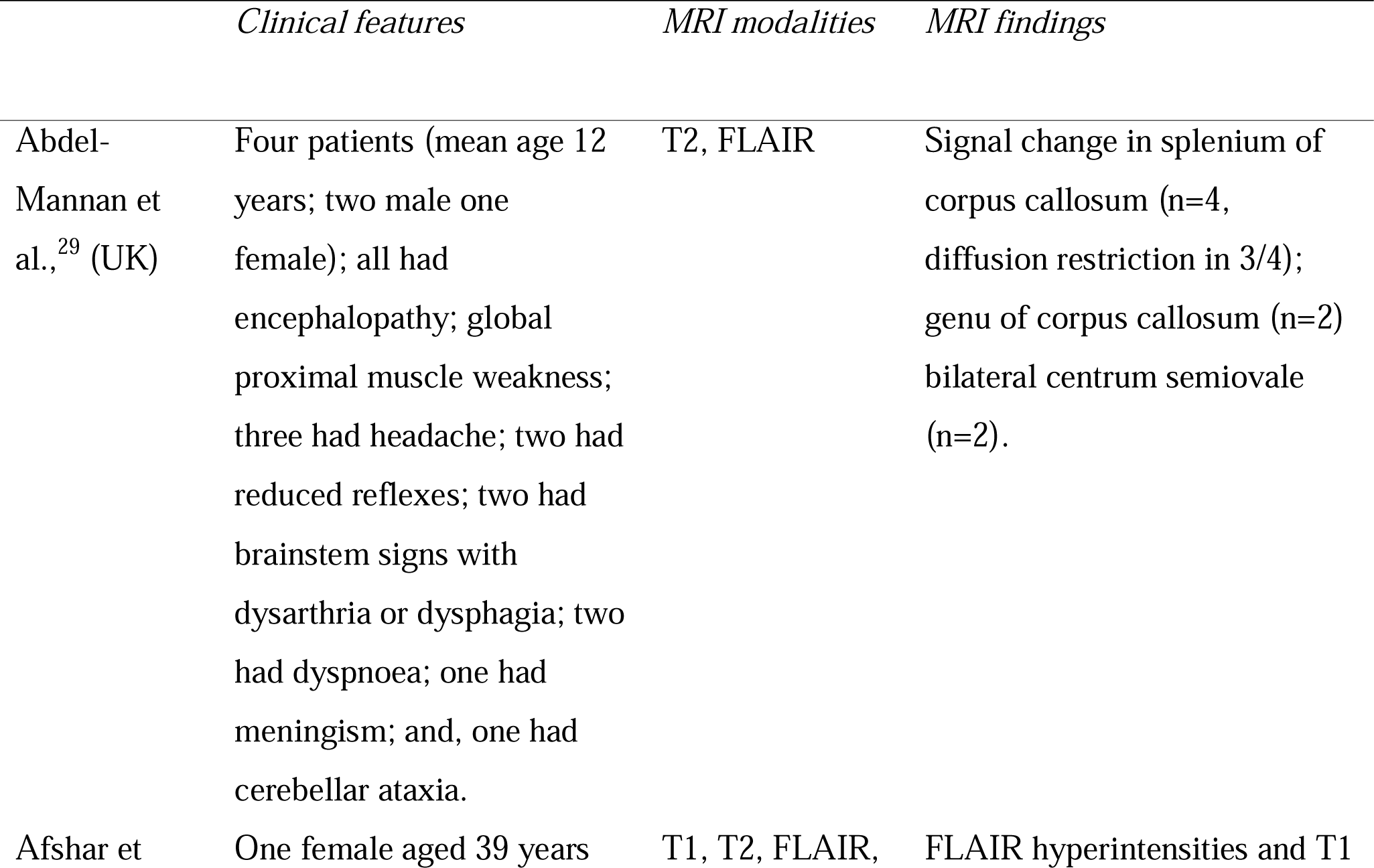
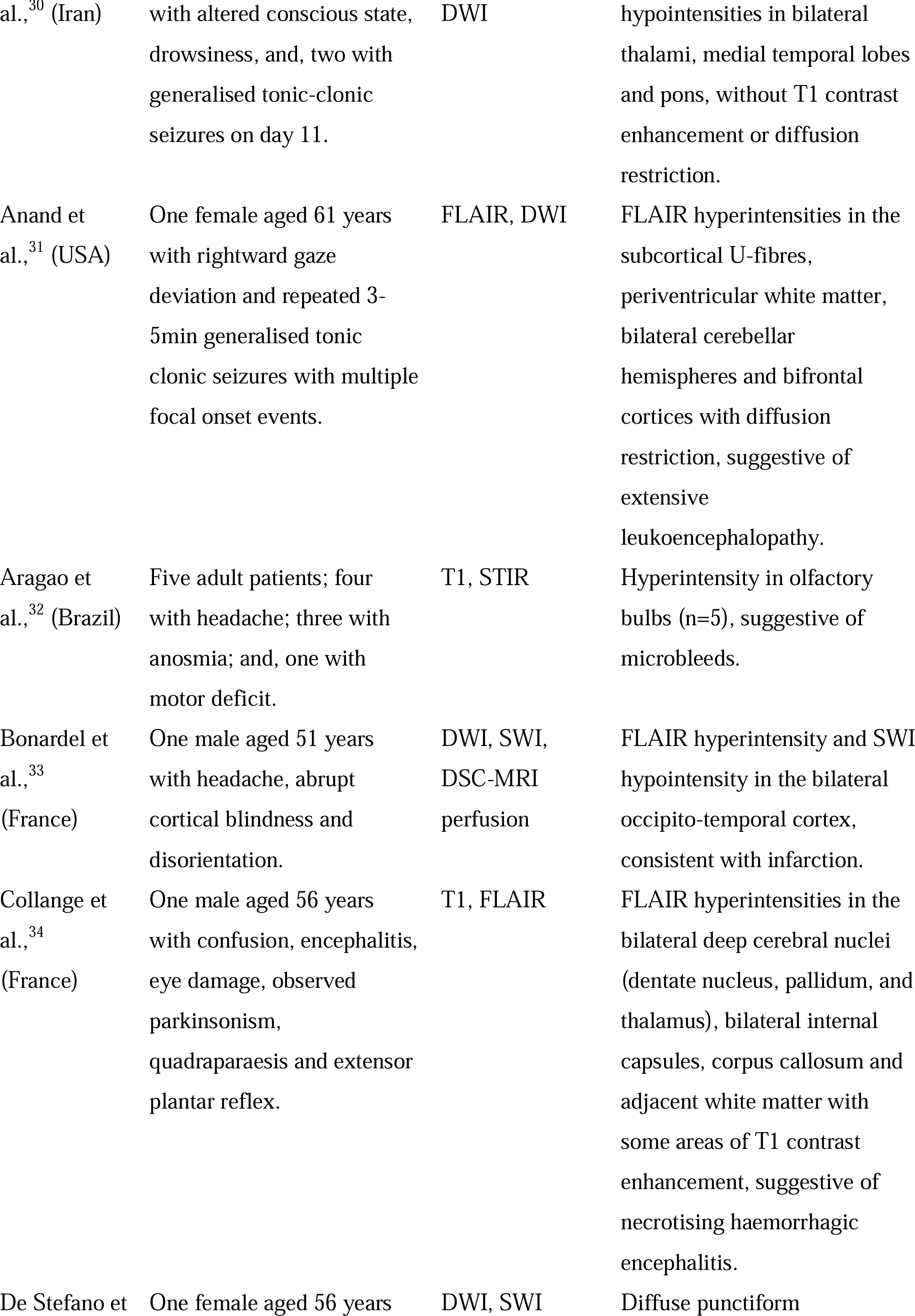
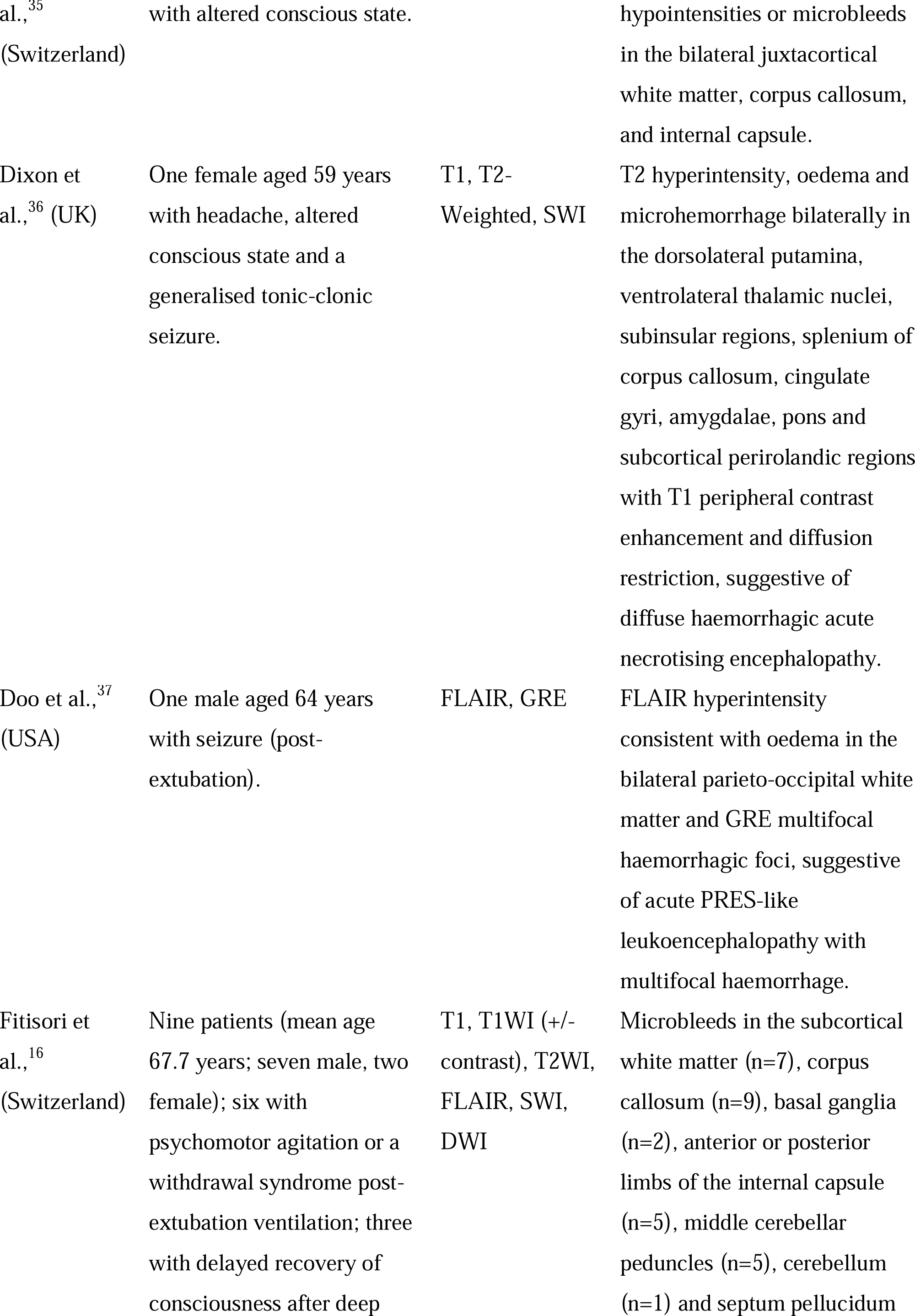
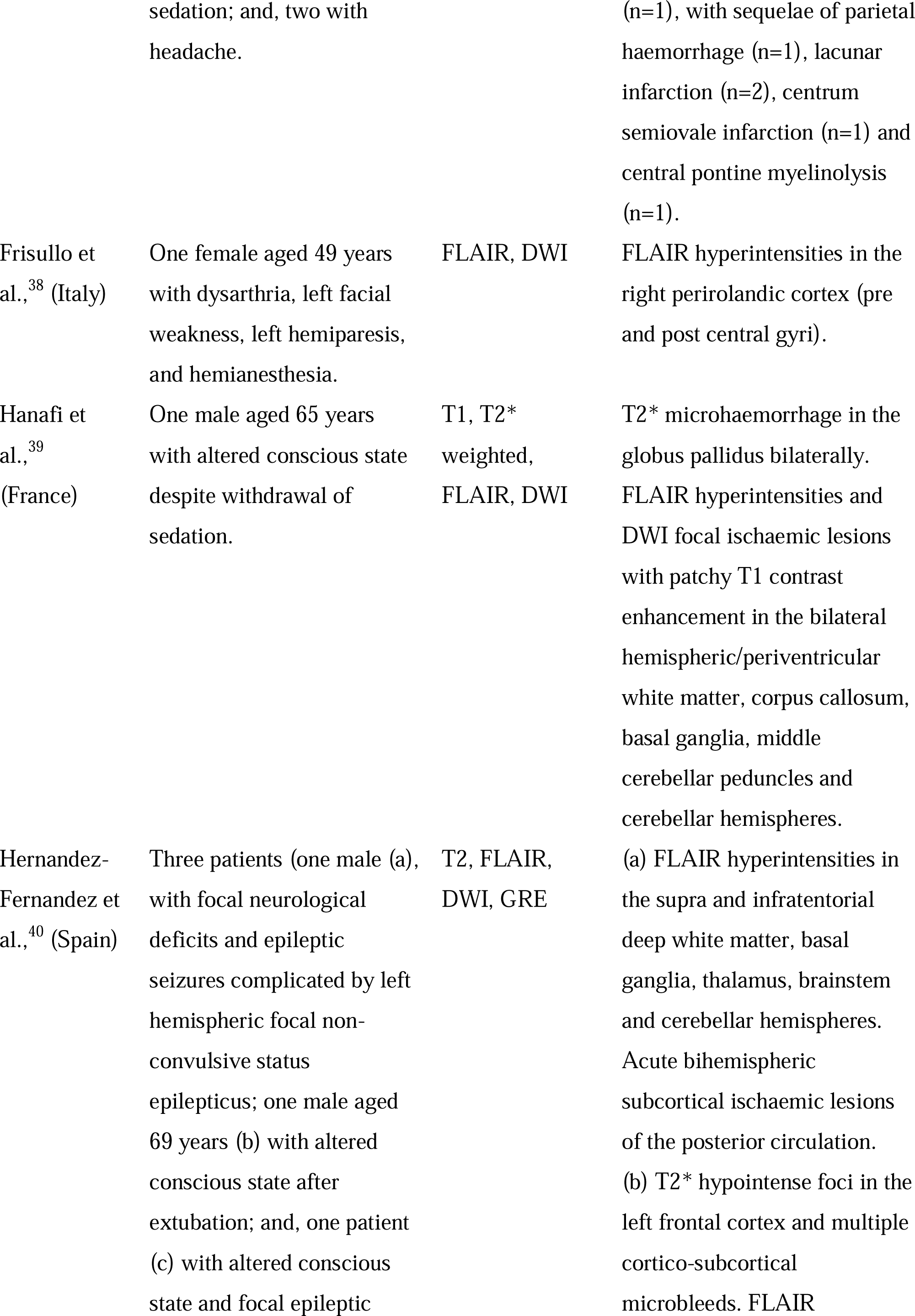
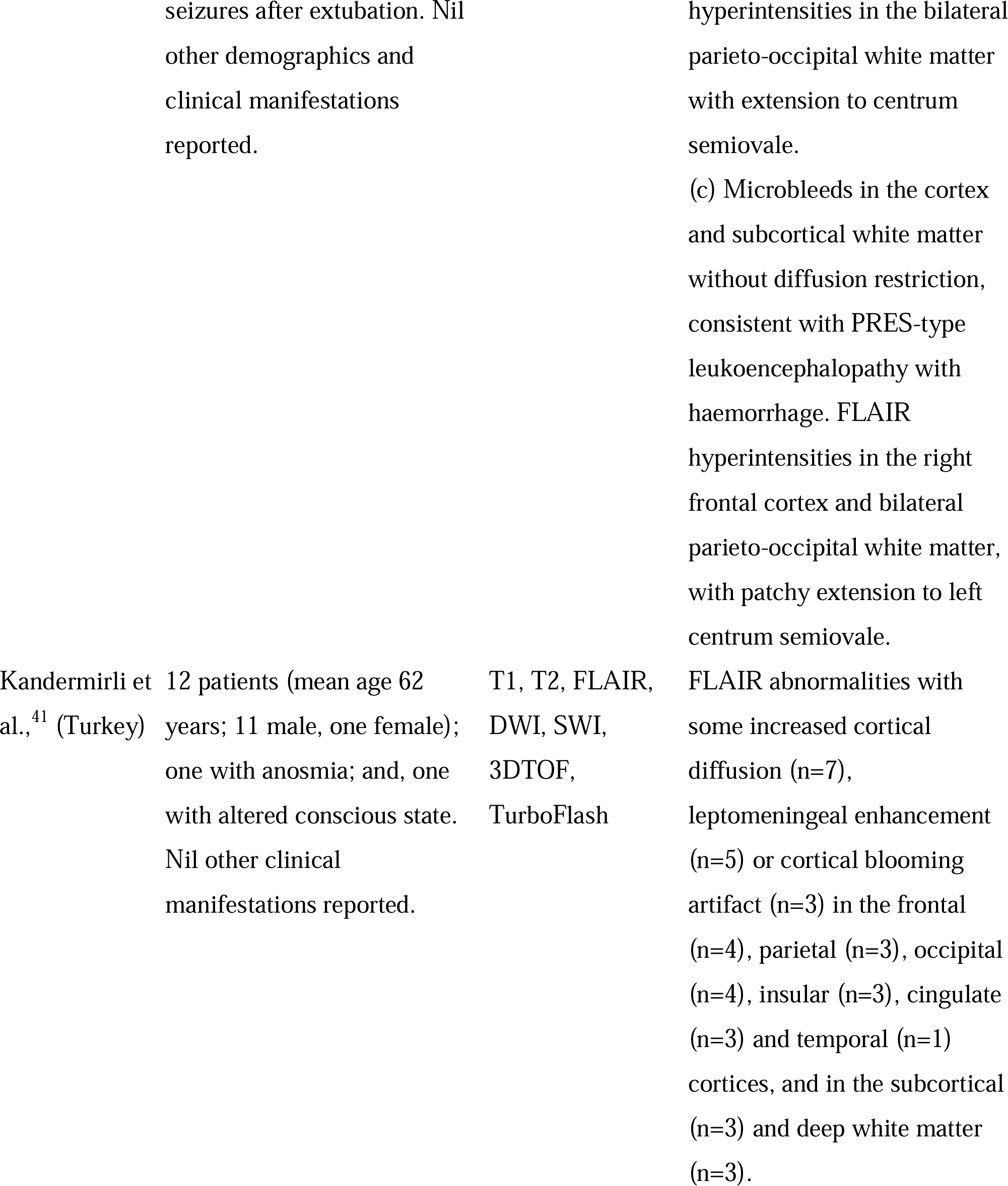
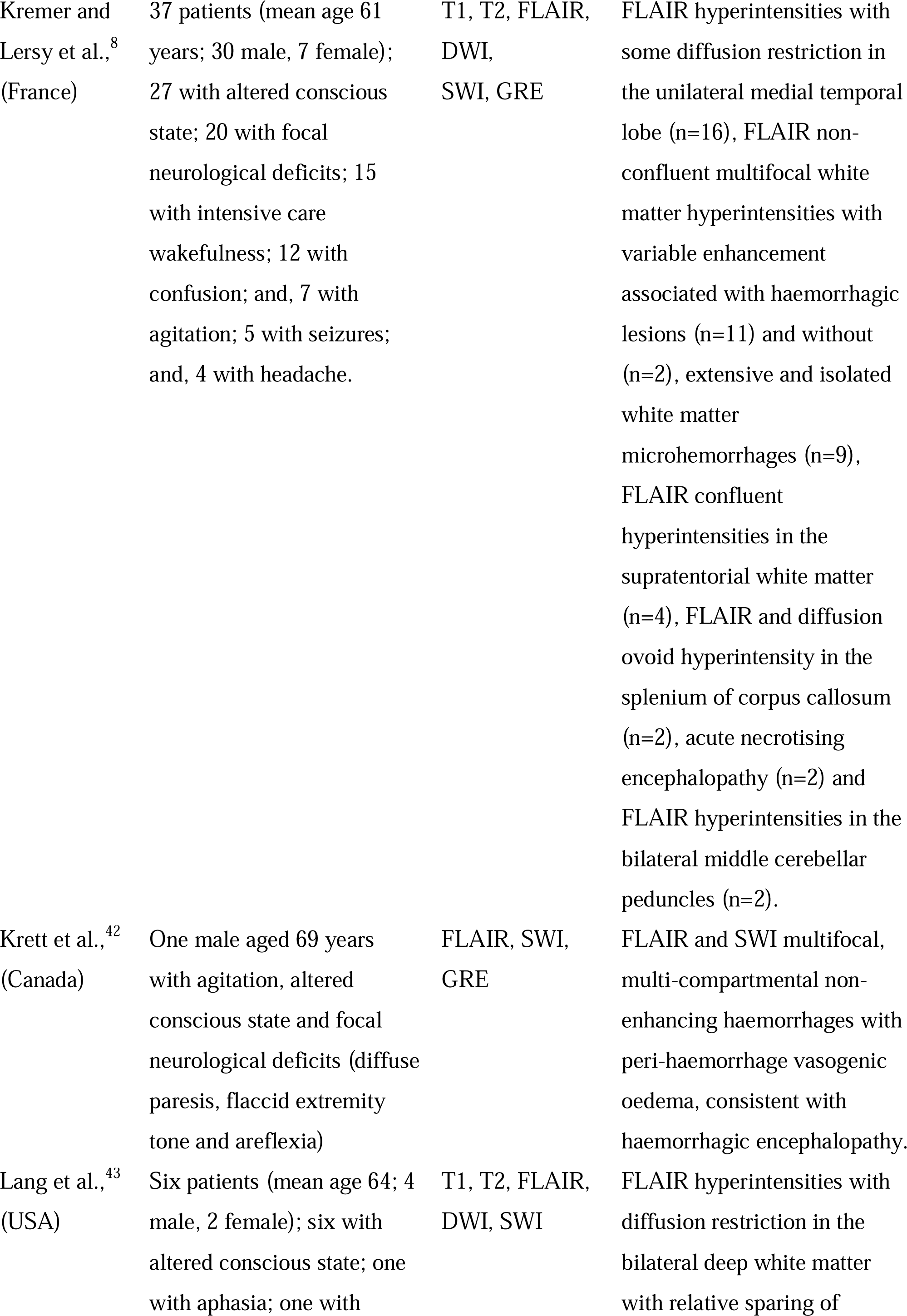
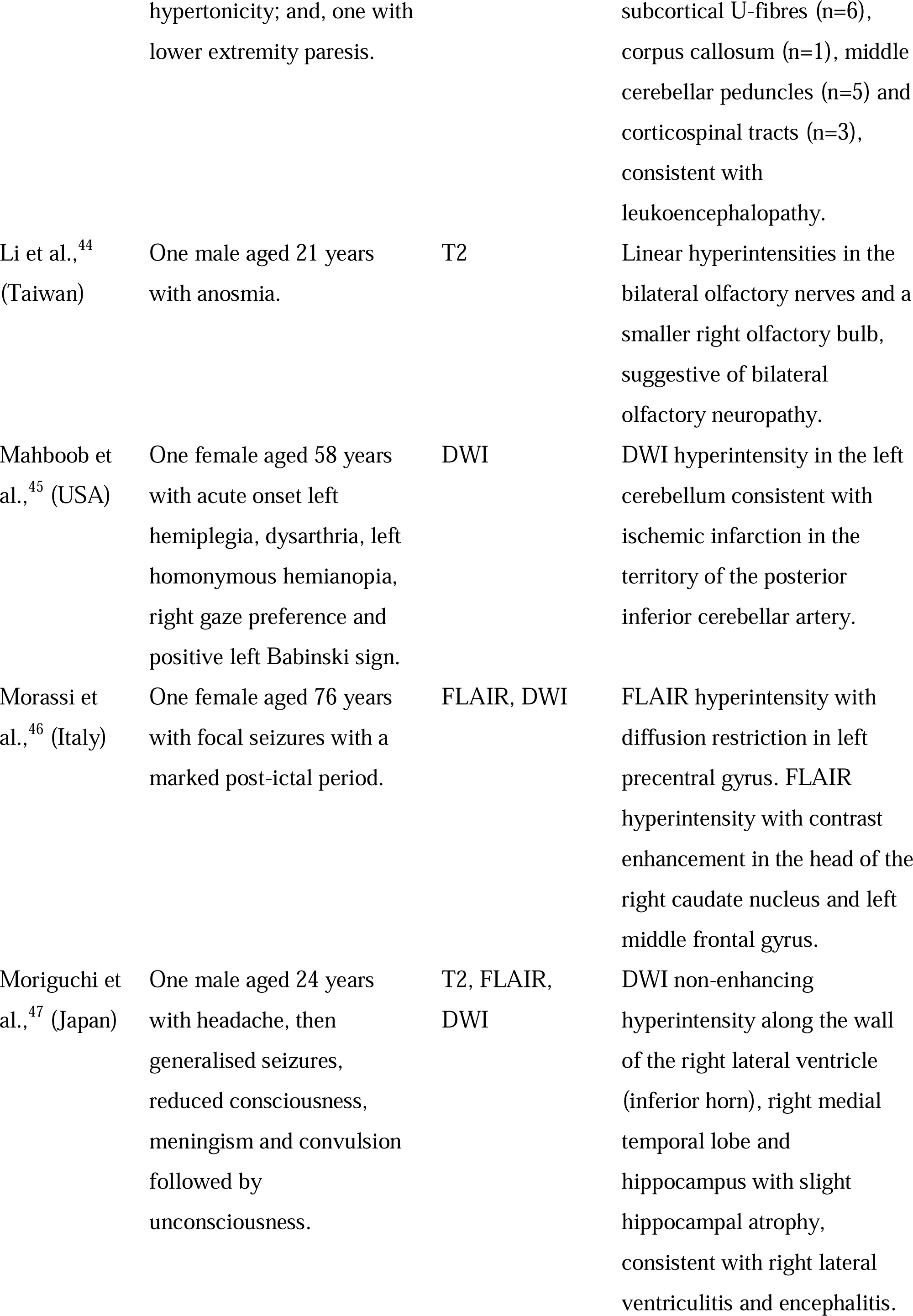
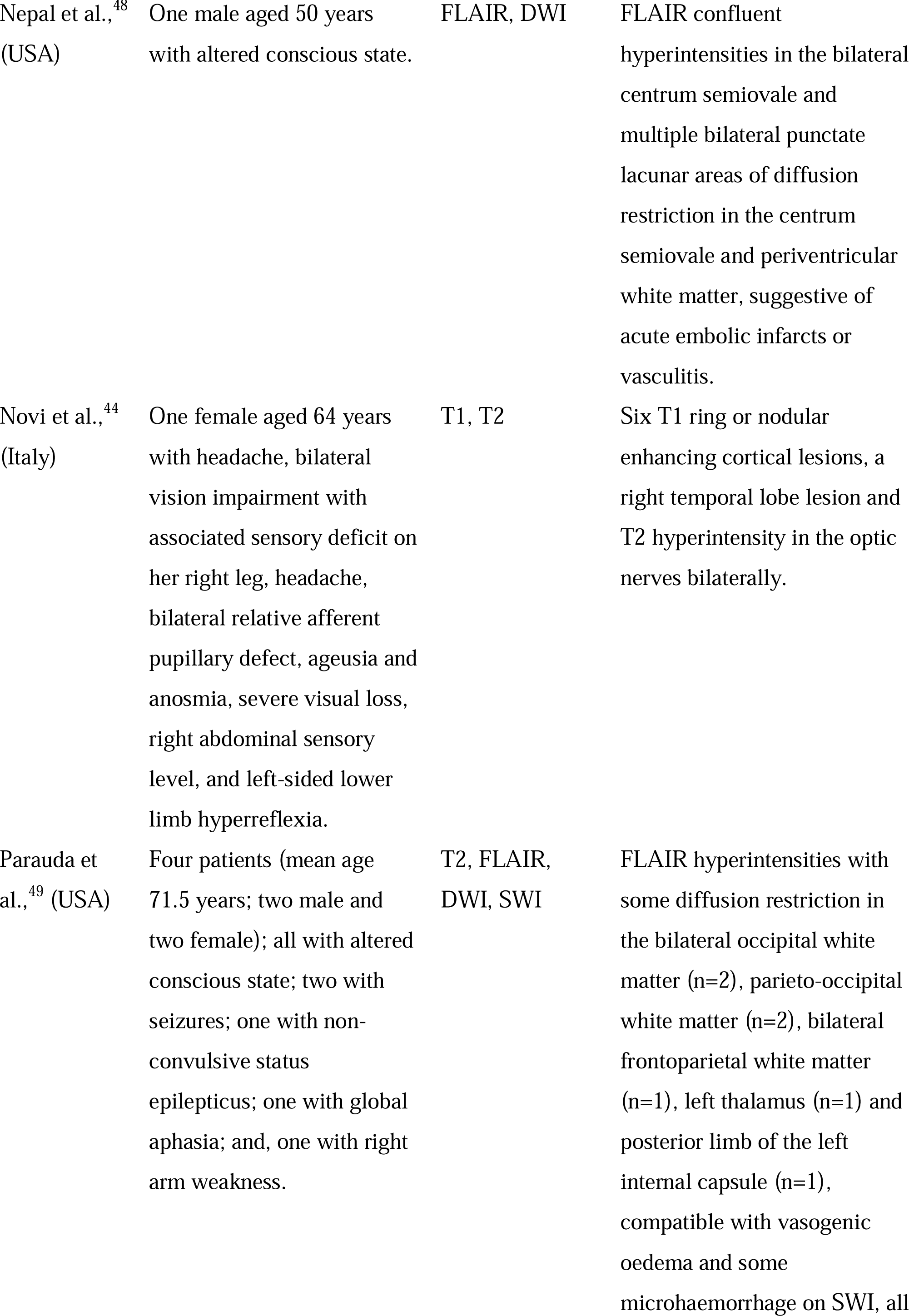
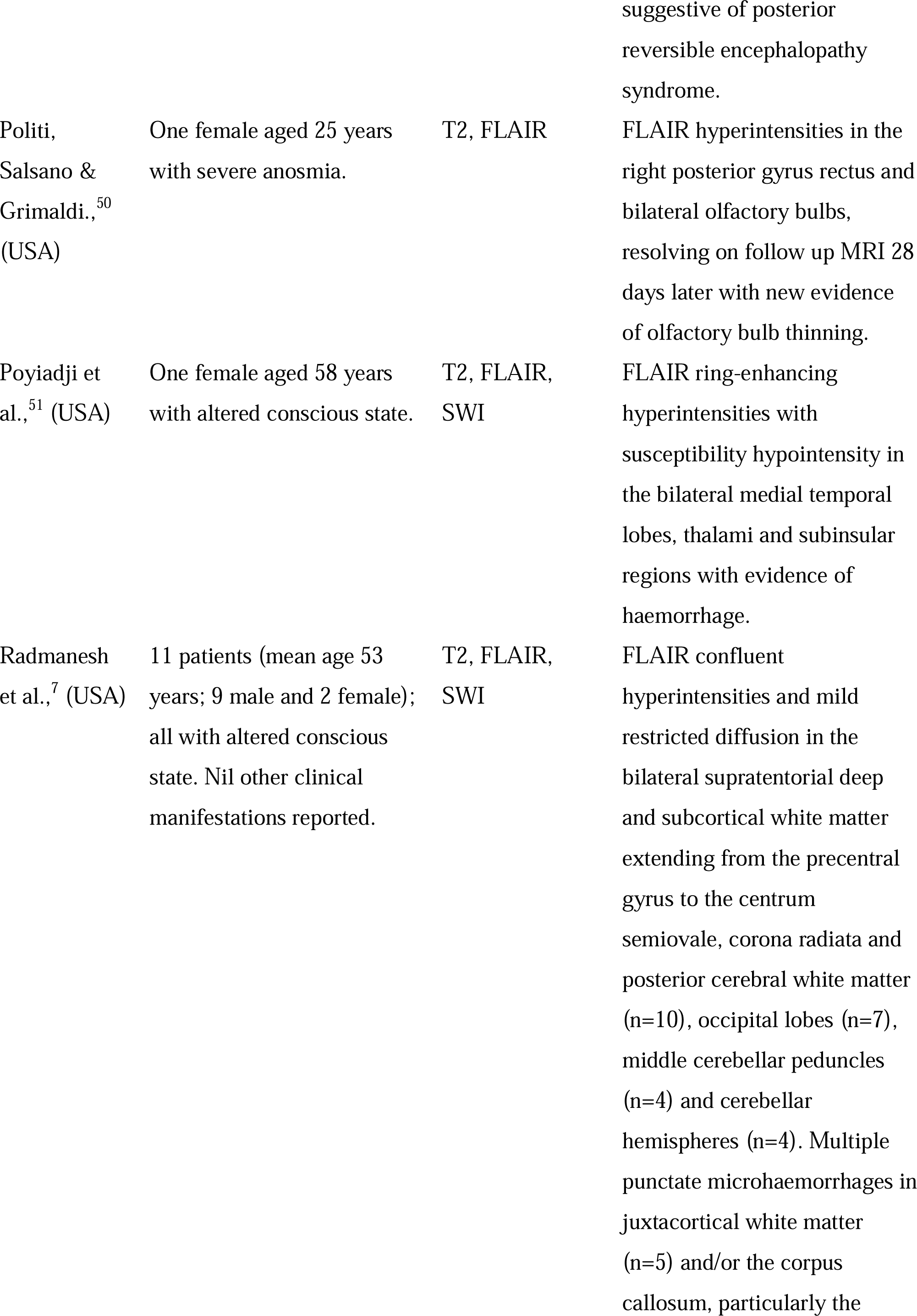
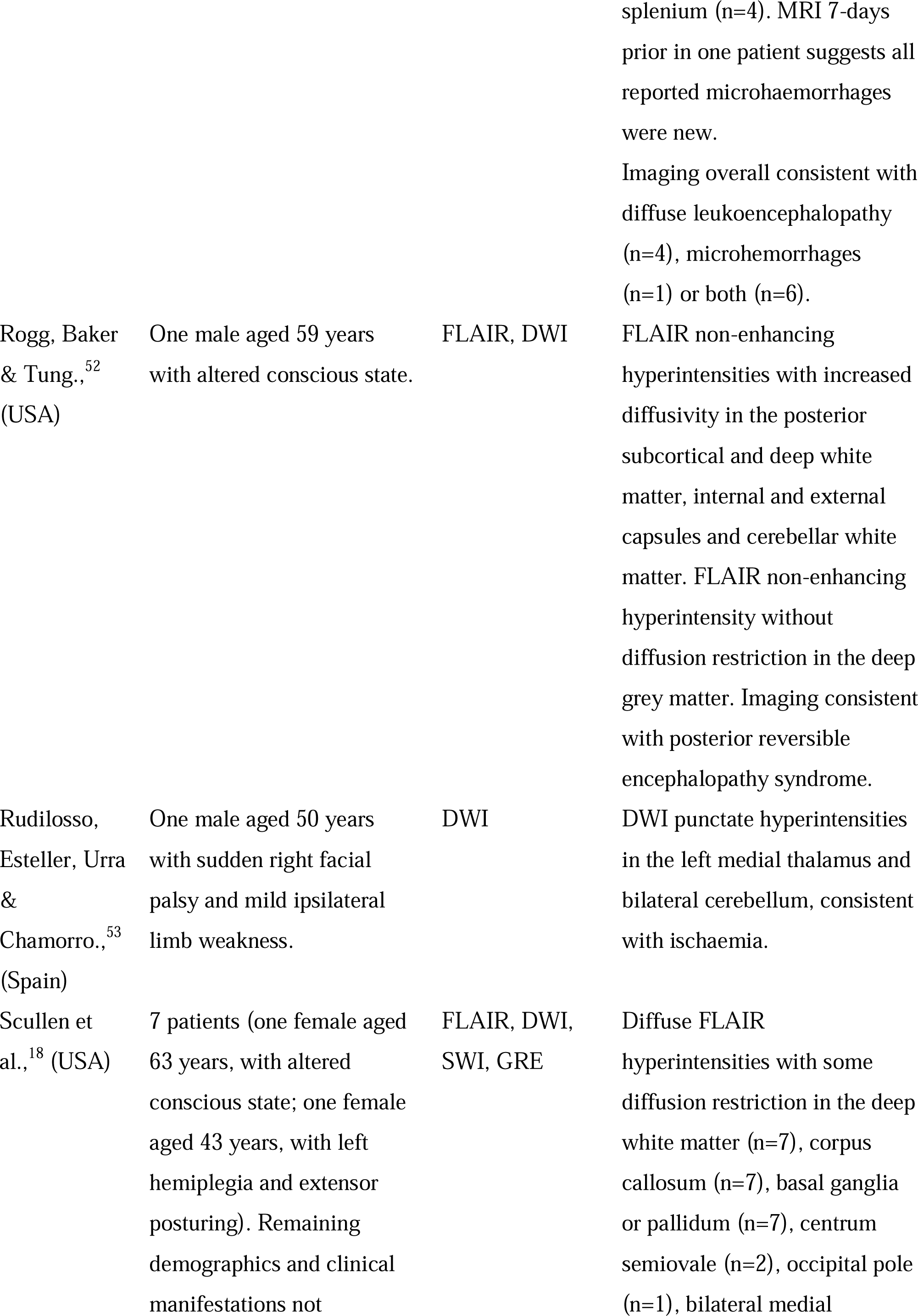
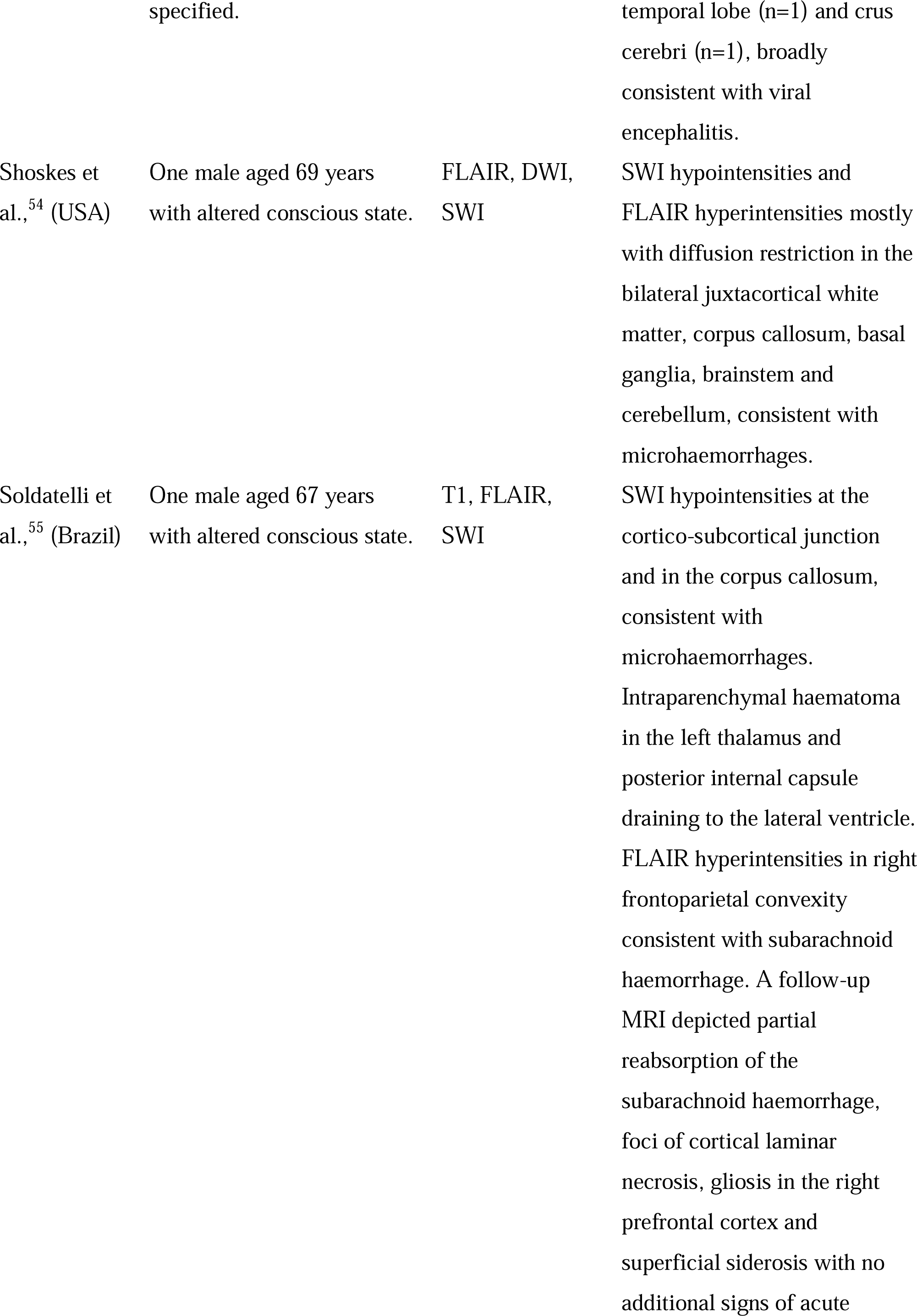
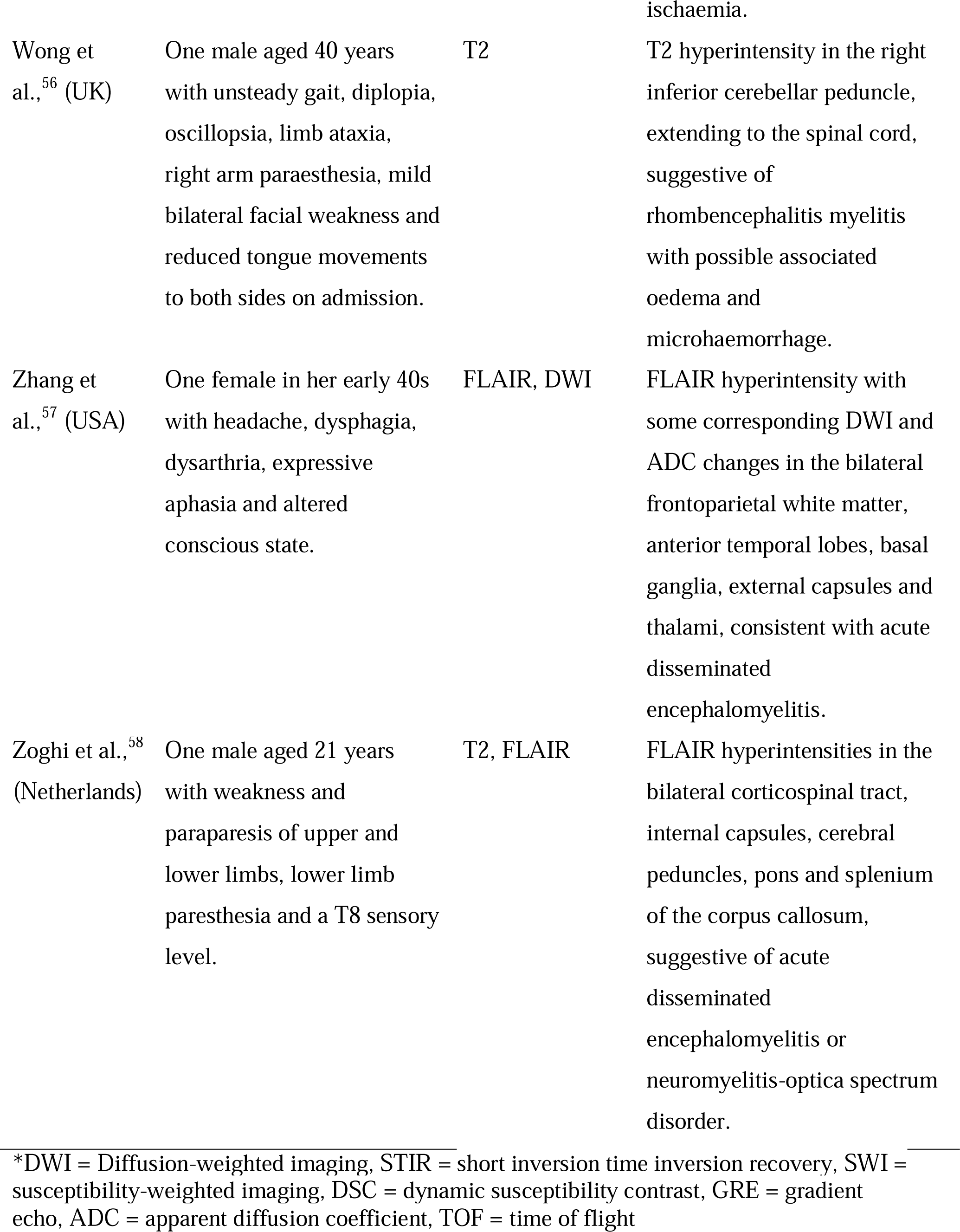
Clinical presentation and MRI findings in COVID-19 patients

### Spatial distribution of neurological events

Figures 1a and b visualise the spatial distribution of white and grey matter neurological events. White matter events were observed within 11 of 42 white matter bundles from the IIT white matter bundles atlas. The highest percentage (26%) of events were observed within the bilateral corticospinal tracts, composed of white matter fibres which connect the primary motor cortex and basal ganglia.^59^. The splenium and middle of the corpus callosum were affected in 14% and 9% of the cases respectively. The remaining tracts show white matter events in less than 9% of cases. Of the cerebral microbleeds observed, a similar pattern emerged; where the largest proportion of cerebral microbleeds were also found in the middle corpus callosum, followed by the splenium of the corpus callosum. Grey matter events were spatially distributed among 41 brain regions within the Desikan-Killiany atlas. The highest proportions (∼10%) of the events were observed in bilateral superior temporal, precentral, and lateral occipital cortices. Sub-cortical events were most frequently identified in the Pallidum.

The highest proportions of events occurred in white matter areas (*a*) such as the corticospinal tract. (*b*) In grey matter, the most vulnerable regions were the bilateral superior temporal cortices, precentral cortices, and pallidum. (*c*) A network diffusion model using structural connectivity edge weights successfully predicted the spread of neurological events. The epicentres of spread which showed the most significant association between predicted and measured distribution of events were the bilateral cerebellum and putamen (*c*).

### Network diffusion model findings

Each of the 84 regions within the IIT Desikan-Killiany grey matter atlas were used as a potential seed for the spread of pathology over time. Figure 1d shows a glass-brain visualisation of the best fit (maximum Pearson’s correlation coefficient value) between empirical events and predicted values determined using NDM. A significant fit was achieved when seeding the spread from the bilateral cerebellum (Pearson’s *r*=0.52, *p* < 0.001 corrected) and putamen (*r*=0.4, *p*=0.02 corrected). Other basal ganglia structures also showed moderate associations (*r*>0.3) but were not significant after correction. The spatial distribution of the fit in all regions reflects the consequence of network spread starting from each of the regions. The correlation-time curve demonstrated the fit between empirical data and the NDM predicted events for all bilateral seeds (42 regions) is shown in Supplementary Figure 1a. The association between empirical events and predicted events was low (*r*<0.2) when Euclidian distance between regions was used as network edges instead of the structural connectivity data (Supplementary Figure 1b).

## Discussion

COVID-19 patients are vulnerable to acute neuropathology, commonly in the form of small neurological and cerebrovascular events. We systematically reviewed articles reporting localised MRI findings in COVID-19 patients and spatially encoded them onto a common grey and white matter atlas. We then investigated whether the spatial distribution of these events follows a cortical or subcortical pattern that can be explained by a linear diffusion-based model of pathological spread. We found the *epicentres* of this spread to be the cerebellum and putamen.

### Neurological events in white and grey matter

White matter events were identified most frequently in the corticospinal tract and corpus callosum. The corticospinal tract is a major white matter pathway connecting critical sub-cortical brain regions such as the basal ganglia and thalamus and as such, facilitates information related to voluntary motor control. As a result, diffuse aberrations in the corticospinal tract are associated with motor symptoms such as tendon reflexes, ankle clonus, and bilateral extensor plantar reflexes, which have been commonly reported in COVID-19 patients.^8,38,42,46,60^ Similarly, the corpus callosum plays an important role in interhemispheric communication; which can result in disconnection syndrome and broad neurocognitive deficits.^65^

In grey matter regions, events were identified most frequently in the temporal and precentral gyrus as well as the bilateral thalamus. Alterations in thalamocortical connectivity can disrupt the regulation of consciousness and arousal.^61,62^ As such, acute events in these regions may explain symptoms such as confusion, disorientation, agitation, and loss of consciousness.^8,30,34,40,51,53,55^ Despite their acute manifestation, the accumulation of neurological events in subcortical structures and consequent disruption to distal cortical regions can increase susceptibility to cognitive impairment and decline—bearing significant ramifications for long-term cognitive prognosis. ^63,64^

### The epicentre and mechanism of spread

We found that the critical epicentres for triggering the widespread neurological events to be the cerebellum and putamen. Given the potential for SARS-CoV-2 neurotropism, this finding is interesting, as neuropathology in the piriform cortex could be caused initially by the introduction of a virus through a direct axonal connection with the olfactory bulb.^65^ However, a speculative interpretation could be that the cerebellum, due to its anatomical proximity to large cerebral arteries may serve as the initial site of neuropathology. SARS-CoV-2 may then travel to deep subcortical structures that are supplied sequentially with blood *after* the cerebellum such as the putamen, and then to cortical sites such as the precentral gyrus, via retrograde transsynaptic transmission by hijacking axonal transport mechanisms.^65-70^ While in transit, direct neuronal or endothelial cell disruption may exacerbate the systemic pathophysiology, facilitating cerebrovascular complications and mixed type I/II respiratory failure.^3,65,69^ However, SARS-CoV-2 has rarely been isolated from CSF samples raising doubt over its neurotropism and direct role in neurological event pathogenesis. ^69,71,72^

Patients with neurological symptoms also presented with anosmia,^32,44,50^ encephalopathy, seizures, and changes to vision including cortical blindness and visual confabulation.^33,44,73^ Alterations in olfaction may therefore have a neurological basis, particularly in light of the identified pathology implicating the olfactory bulb in clinical imaging,^32,44,50^ including the presence of microbleeds among these patients. It is plausible that these symptoms relate closely to the mode of infiltration of SARS-CoV-2, with a potential mechanism being direct injury to the nervous system via ACE2 receptor expression on nerve cells, including the olfactory bulb.^37^

Other proposed pathological mechanisms may explain the distribution of neurological events including neuroinflammatory responses and cytokine and hypoxia-induced injury.^4,18,69^ Emerging evidence is characterising COVID-19 as a vascular disease; a hyperinflammatory response with ensuing cytokine storm and coagulopathy which may synergistically contribute to neurological event pathogenesis.^3,4,18,69,74^ COVID-19-associated coagulopathy occurs proportional to disease severity and leads to treatment-resistant thrombotic and haemorrhagic events; characterised by d-dimer elevation with prothrombin prolongation and thrombocytopenia.^74-77^

Furthermore, cytokine- and hypoxia-induced injury to the corpus callosum, particularly the splenium, has been reported within critical illness including acute respiratory distress syndrome and high-altitude cerebral oedema, potentially contributing to a vulnerability in COVID-19.^17,78-83^ Hypoxia directly induces chemical and hydrostatic endothelial cell disruption, promoting vascular permeability and hence contributing to neurological event pathogenesis.^84^ Relative to the cortex, the thalamus, basal ganglia and deep white matter are poorly perfused due to their watershed end-arterial vascular architecture which could exacerbate their baseline hypoxic vulnerability and ultimately promote subcortical neurological events.^84-88^ While the pathogenesis of white matter hyperintensities remains under debate, roles for hypoxia, immune activation, endothelial cell dysfunction and altered metabolism have been posited; not dissimilar from the neuropathological suggestions within COVID-19.^13-89^

### Limitations

This review has several important limitations. Firstly, we translated neurological events into a standard MRI atlas space using a qualitative method; whereby these pathologies were localised using the radiological description of the location and MRI images where available. While this method is inherently subjective and lacks specificity, we used multiple neuroimaging experts and only included data with specific spatial information or MRI images. As such, the qualitative nature of the translation should be considered with caution while interpreting our findings. Secondly, most of the included articles were cross-sectional case studies and hence cannot directly attribute the observed neuropathology to SARS-CoV-2. Therefore, longitudinal neuroimaging is necessary to directly assess causality. Lastly, some of the neurological events included in our study may be explained by the ageing process; whereby white matter hyperintensities are correlated with age.^13,90^ Hence, findings regarding white matter changes; particularly white matter hyperintensities in the centrum semiovale, should be interpreted with caution.

## Conclusion

Patients with COVID-19 exhibit acute neuropathological and cerebrovascular events. These events occur predominantly in white matter tracts such as the corticospinal tract and corpus callosum, as well in grey matter areas such as the pallidum, putamen, thalamus, and cerebellum. These aberrations likely contribute to altered thalamocortical connectivity and may disrupt the regulation of consciousness and arousal. The accumulation of these events in subcortical structures and the consequent disruption to distal regions, may ultimately increase susceptibility to cognitive impairment and decline—bearing significant long-term cognitive ramifications. Given the prevalence and severity of these manifestations, clinicians should consider having a low threshold for investigating neurological symptoms and monitoring potential long-term *sequelae* in COVID-19 patients.

## Data Availability

Data and code for the work is available at https://github.com/govin2000/covidspread

## Author contributions

Nicholas Parsons: data analysis and interpretation, writing, visualization, original draft preparation, Nasia Outsikas: data extraction, curation original draft preparation, Annie Parish: data extraction, Rebecca Clohesy: data extraction, Nilam Thakkar: data extraction, Fiore D’Aprano: data extraction, interpretation, writing and original draft preparation, Fidel Toomey: data extraction, interpretation, writing and original draft preparation, Shailesh Advani: data extraction, interpretation, original draft preparation, Govinda Poudel: data analysis and interpretation, writing, visualization, original draft preparation.

## Supplementary material

**Fig. 1.**
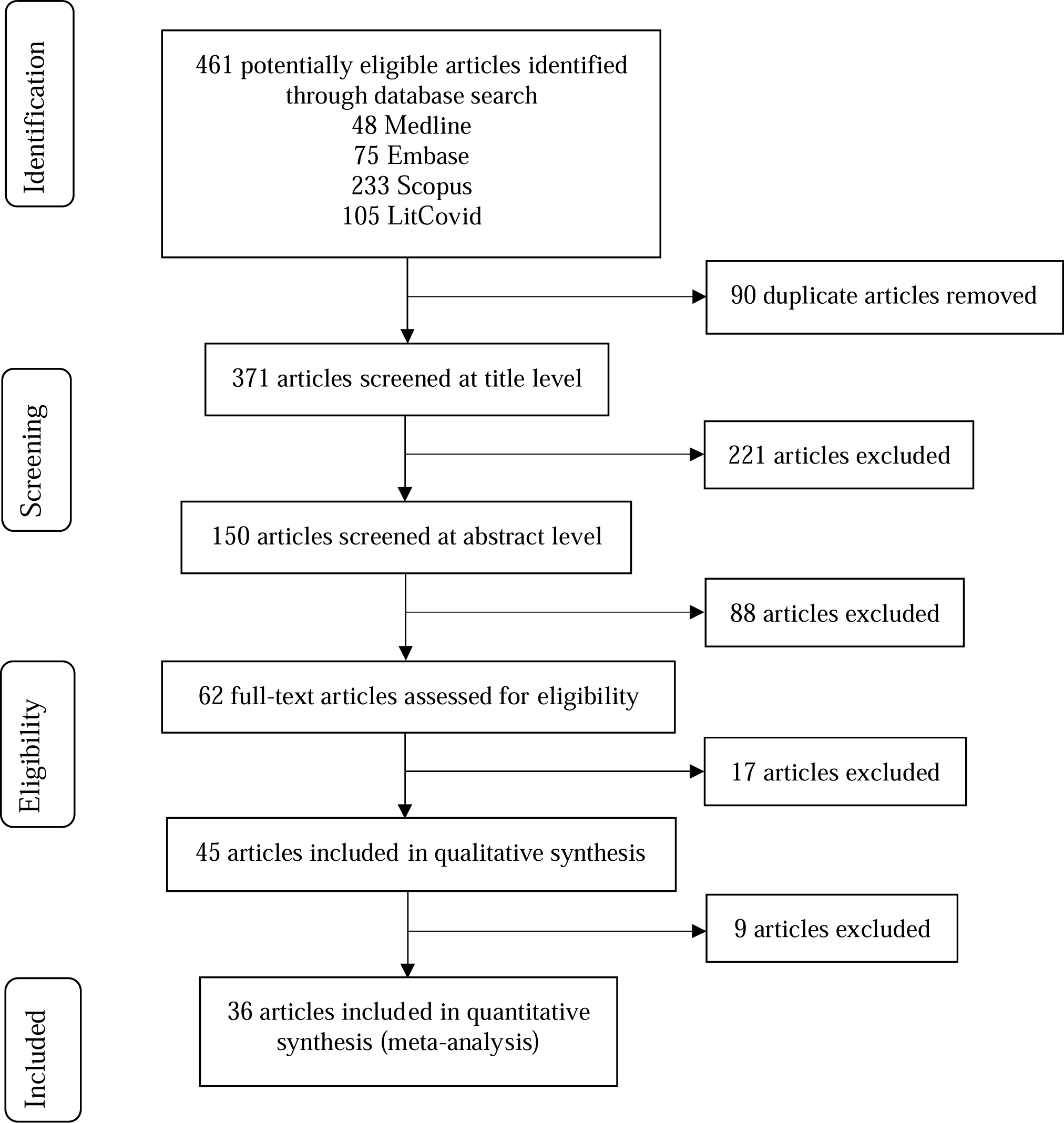
PRISMA flow diagram showing the number of studies screened and included in the meta-analysis.

**Fig 2.**
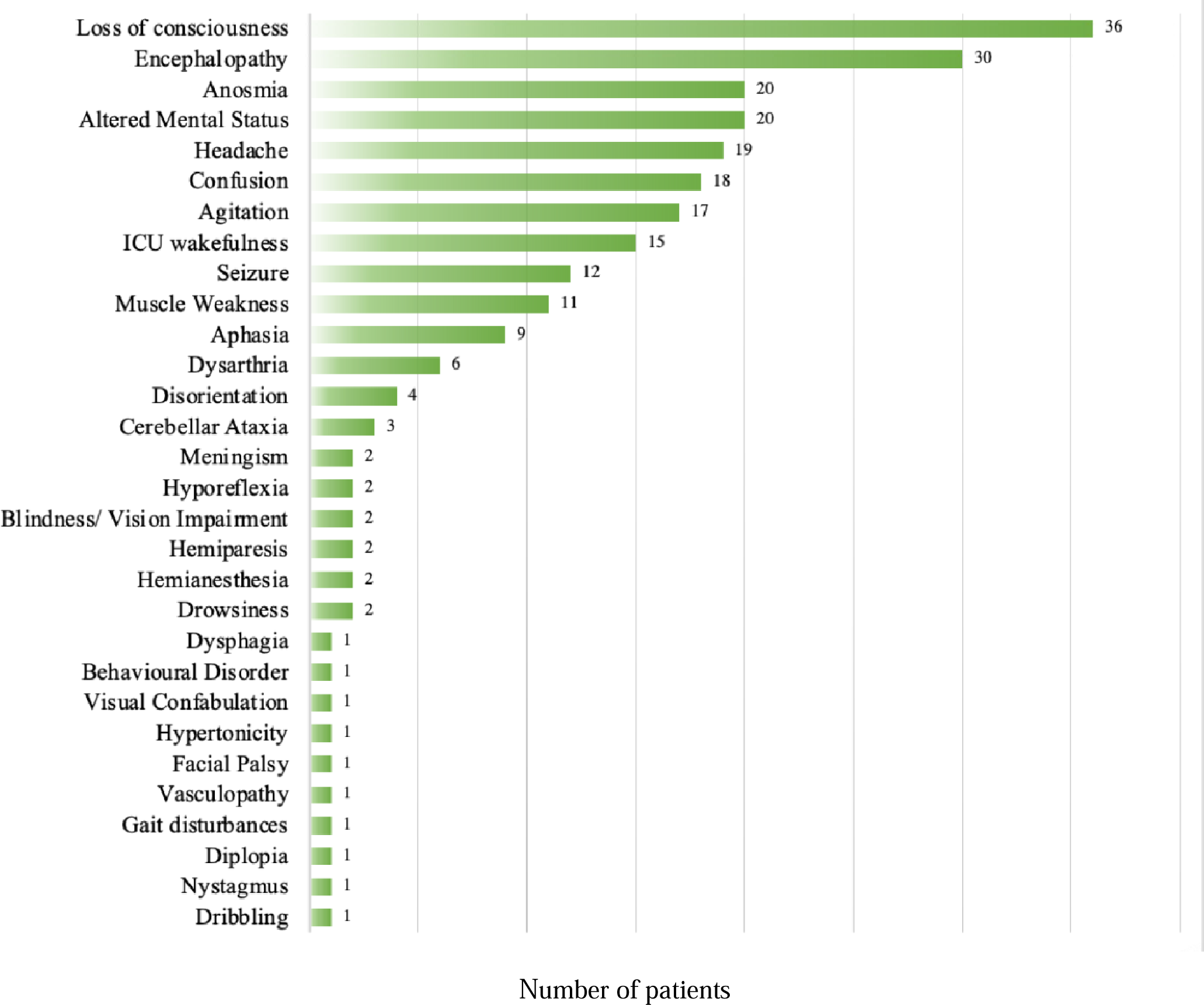
Frequency of COVID-19 relate symptomology.

## Funding disclosure

None

## Conflicts of Interest

None

## Notes

### Competing Interest Statement

The authors have declared no competing interest.

### Clinical Trial

PROSPERO: registration number CRD42020201161

### Author Declarations

Systematic review not requiring ethics approval.

